# Self-reported access to specialty clinics and receipt of health surveillance among U.S. patients with neurofibromatosis 1: A national survey

**DOI:** 10.1101/2024.08.15.24312077

**Authors:** Vanessa L. Merker, Yidan Ma, Lori B. Chibnik, Heather B. Radtke, Kate Kelts, Kaleb Yohay, Nicole J. Ullrich, Scott R. Plotkin, Justin T. Jordan

**Author notes:** **Corresponding Author:** Vanessa L. Merker, Massachusetts General Hospital, 55 Fruit St, Yawkey 9E, Boston, MA 02114. These authors contributed equally to the manuscript (Scott R. Plotkin, Justin T. Jordan).

## Abstract

**Background:** Neurofibromatosis 1 (NF1) is a rare, neurogenetic disorder predisposing individuals to central and peripheral nervous system tumors, as well as a wide range of other physical, neurocognitive, and psychosocial issues. Regular health monitoring throughout the lifespan is needed to identify these heterogenous disease complications, but the degree to which this is implemented in routine clinical practice is unknown. This, this study aimed to investigate U.S. patients’ access to specialized NF1 clinics and receipt of evidence-informed health surveillance.

**Methods:** An online cross-sectional survey was distributed in May 2021 to adults with NF1 and parents of children with NF1 enrolled in the NF Registry (n=6908). Rate of NF1 clinic attendance and self-reported receipt of health surveillance amongst all NF Registry participants was estimated using inverse propensity scores. Differences in these outcomes based on participant demographics were assessed using weighted logistic regression and robust linear regression, respectively.

**Results:** 322 individuals responded (160 adults, 162 parents; 4.7% overall response rate). We estimate that 51.7% of children and 35.6% of adults attend NF1 clinics. Younger children were more likely to attend an NF1 clinic, as were adults living in urban areas, with a college degree or higher, or with a household income ≥$130,000 (all ps<0.05). Completion rates for each individual health surveillance evaluation ranged from 41%-79% for children and 33%-61% for adults. Higher rates of recommended evaluations were reported by both adults and children who attend a specialized NF1 clinic, non-Hispanic White adults, and adults with commercial or Medicare insurance (all ps<0.05).

**Discussion:** Adults with NF1 experience significant sociodemographic disparities in care, and patients of all ages attending NF1 specialty clinics receive more recommended health surveillance. Given the limited access to specialty NF clinics, quality improvement efforts are needed to increase access for underserved adults and improve provision of recommended health surveillance outside specialty clinics.

## Introduction

Neurofibromatosis 1 (NF1) is an autosomal dominant genetic disorder that predisposes individuals to develop multiple benign and malignant nerve sheath tumors and many other physical, neurocognitive, and psychosocial manifestations.^1,2^ Because of the complex, heterogeneous, and age-dependent clinical manifestations of NF1, patients require specialist care and regular health monitoring to identify and treat disease complications. Despite consensus recommendations that people with NF1 be evaluated by, managed by, or have care coordination from specialized neurofibromatosis (NF) clinics^3,4^, access to these clinics is limited in the United States (US). In 2007, the Children’s Tumor Foundation established an NF Clinic Network (NFCN) to identify clinics that provide comprehensive medical care to individuals with NF. By 2015, the 50 NF clinics in this network served an estimated 13.4% of the US NF1 population.^5^ Prior research indicated low rates of NF clinic attendance among people residing in the Southwest and Far West and identified that adults with NF1 experience significant travel barriers to reach high-volume centers compared to children with NF1.^5^ However, non-geographic factors that influence whether US NF1 patients attend specialty NF clinics have not been explored.

Clinical practice resources from the American College of Medical Genetics and Genomics (ACMG) and the American Academy of Pediatrics (AAP),^3,6^ as well as additional expert consensus statements,^7,8^ provide guidance on the range and schedule of recommended health surveillance for people with NF1. Prior research has shown that US-based NF specialists support most of these clinical recommendations, with the majority of surveyed individuals strongly agreeing with 17 published NF1 care recommendations.^9^ To date, there is no data on the degree to which US NF1 patients receive these recommended evaluations in routine clinical practice. Comprehensive data on current rates of adherence to these consensus-based health surveillance recommendations could help researchers, health systems, and disease advocacy organizations prioritize their efforts to improve the quality of NF1 care. The goal of this study was to evaluate self-reported attendance at specialty NF1 clinics and receipt of expert recommended care for US-based NF1 patients as an initial step in monitoring the quality and provision of NF1 care at a national level.

## Methods

### Participant Recruitment

In 2012, the Children’s Tumor Foundation established the NF Registry, an international, web-based platform consisting of participant-entered health information.^10^ Enrollees in the NF Registry who lived in the US, had a self-reported diagnosis of NF1, and had previously consented to share their de-identified health information for research purposes were eligible to participate. Both adults with NF1 (age ≥18) and parents/caregivers of children with NF1 (age <18) were eligible to participate.

Recruitment for the survey ran from May 4 to May 31, 2021, coinciding with NF Awareness Month. Initial survey invitations and up to two follow-up reminders were emailed to all eligible enrollees by the NF Registry administrator. All survey responses were linked to each individual’s NF Registry account, preventing multiple participation from the same participants; however, participants’ identities were not shared with the study team to protect patient privacy and confidentiality.

### Standard Protocol Approvals, Registrations, and Patient Consents

This study was approved by the Western Institutional Review Board. All survey respondents provided voluntary informed consent to participate in the study and there was no compensation for participation.

### Data Collection

This cross-sectional survey was administered online within the NF Registry interface using the OpenApp platform. Respondents were asked to self-report demographic data [e.g., age, gender, race/ethnicity, state of residence, location type (urban/suburban/rural), education level, insurance coverage type, household income] and information about their NF care team [e.g., timing of most recent visit with their NF care team, attendance at any specialized NF clinic within the prior 3 years, and attendance at an NFCN clinic within the prior 3 years]. Participants who had not attended a specialized NF clinic within the last 3 years were asked to rate whether five pre-identified barriers to accessing specialized clinics were a major problem, minor problem, or not at all a problem; these participants also had the option to provide free-text answers describing any additional reasons for not attending an NF clinic.

Participants were also asked to self-report whether they had received various ACMG- and AAP- endorsed health evaluations. Seven evaluations were selected for analysis based on their high rates of agreement among US NF specialists;^9^ suitability for self-report; and applicability to a large proportion of children or adults with NF1 (rather than assessing age-, gender-, or symptom-specific guidelines). These evaluations included blood pressure check, skin exam, scoliosis screening, assessment of pubertal development (pediatric participants only), tracking of developmental milestones/school progress (pediatric participants only), education about warning signs/symptoms of malignant peripheral nerve sheath tumor (MPNST) (adult participants only), and education about family planning options for this genetic disease (adult participants only). Most survey questions assessed care provided at the most recent visit or within the prior year, in accordance with the recommended frequency in published guidance.

### Data Analysis

To explore how representative our survey sample was, we compared the demographics of survey respondents to all living, US-based NF Registry members with NF1 who had provided, at a minimum, their age and gender. Differences in race/ethnicity were assessed using mutually exclusive categories established by the NF Registry. Geographic differences were assessed by aggregating state-level residence data into 8 US regions as defined by the Bureau of Economic Analysis.^11^ Differences in clinic type were assessed according to clinics’ membership in the NFCN at the time of the survey.

Due to significant differences in survey respondents and non-respondents in age, geographic region, and attendance at NFCN clinics [Supplemental Table 1], we utilized analysis techniques that allow for re-weighting of participants based on these three characteristics to minimize non-response error and provide more generalizable estimates. We used inverse propensity score weighting to estimate the rate of specialized NF clinic attendance, barriers to NF clinic attendance, and receipt of recommended health surveillance among all people with NF1 enrolled in the NF registry. To examine factors affecting attendance at NF clinics, we used 1) weighted logistic regression to examine potential sociodemographic disparities in clinic attendance and 2) qualitative content analysis to inductively summarize survey respondents’ free-text comments describing barriers to attending a specialized NF clinic.^12^

We also examined whether participant demographics and location/timing of NF care impacted receipt of recommended health surveillance using robust linear regression [regression with an iteratively reweighted least squares which allows for the inverse propensity score weighting and is less sensitive to divergence from the normality assumption]. Our primary outcome measure for receipt of recommended care was the number of recommended NF-related evaluations (ranging from 0-5) completed per individual, analyzed separately for children and adults with NF1. For adults, the five evaluations were annual blood pressure check, skin exam, scoliosis screening, malignant peripheral nerve sheath tumor (MPNST) education, and family planning education; for children, the five evaluations were annual blood pressure check, skin exam, scoliosis screening, assessment of pubertal development, and tracking of developmental milestones/school progress. Given the risk of type 1 error associated with assessing multiple predictors of receiving multiple evaluations, we did not examine how participant demographics or NF1 care location/timing impacted receipt of each individual evaluation criteria. Data was analyzed in R program and a p-value of ≤0.05 was considered statistically significant for all tests.

### Data Availability

De-identified participant data may be requested from the NF Registry by emailing. All requests are reviewed by the NF Registry Data Access Committee to ensure appropriate scientific and ethical use of the data.

## Results

A total of 322 US-based NF1 patients and parents/caregivers completed the survey, representing 4.7% (322/6908) of eligible NF Registry enrollees. There were no significant differences in gender or race/ethnicity between the subset of survey respondents and all NF Registry enrollees; significant differences in age, U.S. region, and NFCN clinic attendance were adjusted for using inverse propensity scores (Supplemental Table 1). Survey respondent demographics and location/timing of NF1 care are shown in Table 1. The majority of respondents (83.3% of children and 55.0% of adults) had seen their NF1 care provider in the prior year, and 14.0% of respondents had delayed or foregone necessary medical care due to cost in the prior year.

**Table 1.** Survey Respondent Characteristics by Age Group.

|  | Children<br>(n=162) | Adults<br>(n=160) |
| --- | --- | --- |
| Age, mean (SD) | 7.5 years (4.5) | 43.8 years<br>(14.7) |
| Gender, n (%) |  |  |
| Female | 75 (46.3%) | 112 (70%) |
| Male | 87 (53.7%) | 48 (30%) |
| Race/Ethnicity, n (%) |  |  |
| White | 126 (77.8%) | 126 (78.8%) |
| Native American or Alaskan Native | 2 (1.2%) | 5 (3.1%) |
| Black, Afro-Caribbean, or African-American | 9 (5.6%) | 9 (5.6%) |
| Native Hawaiian or other Pacific Islander | 0 (0.0%) | 0 (0.0%) |
| Latino/Hispanic | 9 (5.6%) | 4 (2.5%) |
| East Asian | 5 (3.1%) | 6 (3.8%) |
| South Asian | 3 (1.9%) | 2 (1.3%) |
| Other | 8 (4.9%) | 8 (5.0%) |
| US Region, n (%) |  |  |
| Far West | 40 (24.7%) | 20 (12.5%) |
| Rocky Mountain | 11 (6.8%) | 11 (6.9%) |
| Southwest | 15 (9.3%) | 16 (10.0%) |
| Plains | 8 (4.9%) | 12 (7.5%) |
| Great Lakes | 24 (14.8%) | 16 (10.0%) |
| Southeast | 25 (15.4%) | 28 (17.5%) |
| Mid-Atlantic | 32 (19.8%) | 37 (23.1%) |
| New England | 6 (3.7%) | 18 (11.3%) |
| Geographic Location, n (%) |  |  |
| Rural | 30 (18.5%) | 25 (15.6%) |
| Suburban | 100 (61.7%) | 92 (57.5%) |
| Urban | 27 (16.7%) | 40 (25.0%) |
| Not Sure | 1 (0.6%) | 2 (1.3%) |
| Other | 0 (0.0%) | 1 (0.6%) |
| Missing | 4 (2.5%) | 0 (0.0%) |
| Primary Insurance Coverage, n (%) |  |  |
| Commercial or Other Insurance | 126 (77.8%) | 124 (77.5%) |
| Medicaid | 32 (19.8%) | 13 (8.1%) |
| Medicare | 3 (1.9%) | 22 (13.8%) |
| None | 0 (0%) | 1 (0.6%) |
| Missing | 1 (0.6%) | 0 (0%) |
| Family Income, n (%) |  |  |
| Less than \$25,000 | 15 (9.3%) | 31 (19.4%) |
| \$25,000 – \$49,999 | 15 (9.3%) | 37 (23.1%) |
| \$50,000 – \$79,999 | 21 (13.0%) | 28 (17.5%) |
| \$80,000 – \$129,999 | 39 (24.1%) | 23 (14.4%) |
| \$130,000 – \$250,000 | 45 (27.8%) | 11 (6.9%) |
| More than \$250,000 | 19 (11.7%) | 3 (1.9%) |
| Prefer not to say | 7 (4.3%) | 26 (16.3%) |
| Missing | 1 (0.6%) | 1 (0.6%) |
| Education, n (%) |  |  |
| Some high school | N/A | 6 (3.8%) |
| Completed high school |  | 31 (19.4%) |
| Some college |  | 29 (18.1%) |
| Completed an associate's (2 year) degree |  | 18 (11.3%) |
| Completed a bachelor's (4 year) degree |  | 39 (24.4%) |
| Completed graduate level degree |  | 32 (20.0%) |
| Missing |  | 5 (3.1%) |
| Primary Language, n (%) |  |  |
| English | 157 (96.9%) | 157 (98.1%) |
| Spanish | 5 (3.1%) | 3 (1.9%) |
| Other | 0 (0.0%) | 0 (0.0%) |
| Missing | 0 (0.0%) | 0 (0.0%) |
| Attendance at NFCN clinic, n (%) |  |  |
| Yes | 120 (74.1%) | 79 (49.4%) |
| No | 38 (23.4%) | 47 (29.3%) |
| Missing | 4 (2.5%) | 34 (21.3%) |
| Last Visit to NF Care Provider, n (%) |  |  |
| Within the last one year | 135 (83.3%) | 88 (55.0%) |
| One to three years ago | 17 (10.5%) | 24 (15.0%) |
| More than three years ago | 1 (0.6%) | 21 (13.1%) |
| Not sure | 2 (1.2%) | 23 (14.4%) |
| Missing | 7 (4.3%) | 4 (2.5%) |

### Attendance at NF Specialty Clinics

We estimate that 51.7% of children and 35.6% of adults in the NF Registry visited a specialty NF clinic within the prior 3 years. Adults living in urban areas, with a 2-year college degree or higher, or with a household income ≥$130,000 were more likely to have visited a specialized NF clinic in the prior 3 years (all ps <0.05); age, gender, ancestry, and insurance coverage were not significantly related to NF clinic attendance (Table 2). The largest predictor of adult NF clinical attendance was household income; adults with the highest income (>$130,000, approximately the top quintile of U.S. household income) had 12.9 times higher odds of attending a specialized NF clinic than adults with the lowest income (<$25,000, approximately the lowest quintile of U.S. household income). For children, only age was significantly associated with NF clinic attendance; for each additional year of age, there was 11% decreased odds that a child had visited a specialized NF clinic in the prior 3 years (p<0.001).

**Table 2:** Factors Affecting Attendance at an NF Specialty Clinic in the Prior 3 Years.

|  | Children odds ratio<br>(p-value) | Adult odds ratio<br>(p-value) |
| --- | --- | --- |
| Age (years) | **0.89 (p < 0.001)** | 0.98 (p = 0.10) |
| Gender |  |  |
| Male | *reference* | *reference* |
| Female | 0.61 (p = 0.14) | 0.73 (p = 0.39) |
| Race/Ethnicity |  |  |
| Non-Hispanic White | *reference* | *reference* |
| Black | 0.18 (p = 0.08) | 1.00 (p = 1.00) |
| Other<sup>a</sup> | 1.10 (p = 0.85) | 0.75 (p = 0.56) |
| Household Income |  |  |
| <\$25,000 | *reference* | *reference* |
| \$25,000-\$129,999 | 0.84 (p = 0.79) | 1.89 (p = 0.21) |
| >\$130,000 | 0.50 (p = 0.27) | **12.93 (p = 0.004)** |
| Primary Medical Insurance |  |  |
| Medicaid | *reference* | *reference* |
| Medicare | 0.65 (p = 0.70) | 1.54 (p = 0.60) |
| Commercial or Other Insurance | 1.11 (p = 0.82) | 2.90 (p = 0.13) |
| Home Location |  |  |
| Rural | *reference* | *reference* |
| Suburb | 1.14 (p = 0.74) | 2.07 (p = 0.23) |
| Urban | 1.67 (p = 0.41) | **3.77 (p = 0.04)** |
| Education |  |  |
| No college degree | N/A | *reference* |
| College degree or higher<sup>b</sup> |  | **3.03 (p = 0.003)** |

Note: Odds ratios and p-values were calculated from weighted logistic regressions adjusted for age. All p-values <0.05 are bolded. <sup>a</sup>Includes Latino/Hispanic, Asian, Native American or Alaskan Native, Native Hawaiian or other Pacific Islander, and Other due to small sample sizes. <sup>b</sup>Includes 2- and 4-year undergraduate degrees and all graduate/professional degrees.

Participants who did not attend a specialized NF clinic rated the importance of five pre-identified barriers to visiting these clinics; responses were then weighted using inverse propensity scores (Supplemental Table 2). The most frequently endorsed “major problem” to clinic attendance was insurance coverage for adults (18.5%) and lack of knowledge that specialized NF clinics exist for parents of children with NF1 (17.0%). In addition, 68 of 121 (56.2%) of these respondents provided free-text comments regarding barriers to NF clinic attendance. The most common barrier was the travel required due to the lack of nearby NF clinics (n=19). The second most frequent barrier was perceived lack of value in attending a specialized clinic (n=18). These individuals fell into three categories: 1) they already had a doctor or team of doctors they felt could follow their NF adequately, e.g., “His specialist works within a division of neurology at a major hospital, and we like him a lot”; 2) they reported mild symptoms that did not seem to require specialized care, e.g., “Not needed. Very mild case. Does not affect my day-to-day life”, or 3) they were unclear about the added benefit of a specialized clinic, e.g., “I don’t know if there is anything a specialized NF can offer me”. Additional recurring barriers included insurance problems/high cost (n=9), lack of knowledge that specialized clinics existed or where they were located (n=7), appointment availability/scheduling difficulty (n=3), and no referral by their primary care provider (n=2).

### Receipt of Evidence-Informed NF1 Clinical Care

We estimate that the proportion of children with NF1 in the NF Registry receiving recommended health surveillance in the prior year was 79% for blood pressure measurement, 69% for tracking of developmental milestones/school progress, 69% for scoliosis screening, 67% for skin exam, and 41% for assessment of pubertal development. The proportion of adults receiving recommended evaluations was 62% for blood pressure measurement, 61% for education about family planning options, 39% for scoliosis screening, 36% for education about MPNST warning signs, and 33% for skin exam. Only 23% of children and 8% of adults received all five recommended evaluations noted above; one quarter of adults and just over half of children received at least 4 out 5 recommended evaluations (Table 3).

**Table 3.** Inverse propensity score weighted estimates of the cumulative number of recommended health surveillance evaluations received by US-based NF Registry participants with NF1.

| Number of<br>Recommend<br>Evaluations Received* | Adults | Children |
| --- | --- | --- |
| 0 | 8.7% | 11.8% |
| 1 | 24.6% | 2.2% |
| 2 | 26.5% | 12.7% |
| 3 | 15.0% | 19.8% |
| 4 | 17.0% | 30.5% |
| 5 | 8.2% | 23.1% |

Age, visiting a member of one’s self-described NF care team in the past year, and visiting an NF specialty clinic in the past 3 years and were significantly associated with the total number of recommended evaluations received for both children and adults (all ps<0.05, Table 4). Teenagers and young adults received the greatest proportion of recommended health surveillance. Patients who attended a specialized NF clinic in the previous 3 years received 0.8 to 0.9 more recommended NF1-related evaluations, on average, than those who had not visited a specialized NF clinic (Figure 1). Race/ethnicity and insurance coverage were significantly associated with the total number of recommended evaluations received for adults only (all ps<0.05). Non-Hispanic White adults received more recommended evaluations than adults of other races/ethnicities; for example, this group received almost a full evaluation more than Black adults on average. Adults with commercial insurance or Medicare received at least one more evaluation, on average, than adults with Medicaid. Gender, family income, and location (urban/suburban/rural) were not statistically significantly associated with the number of evaluations received for either adults or children.

**Figure 1.**
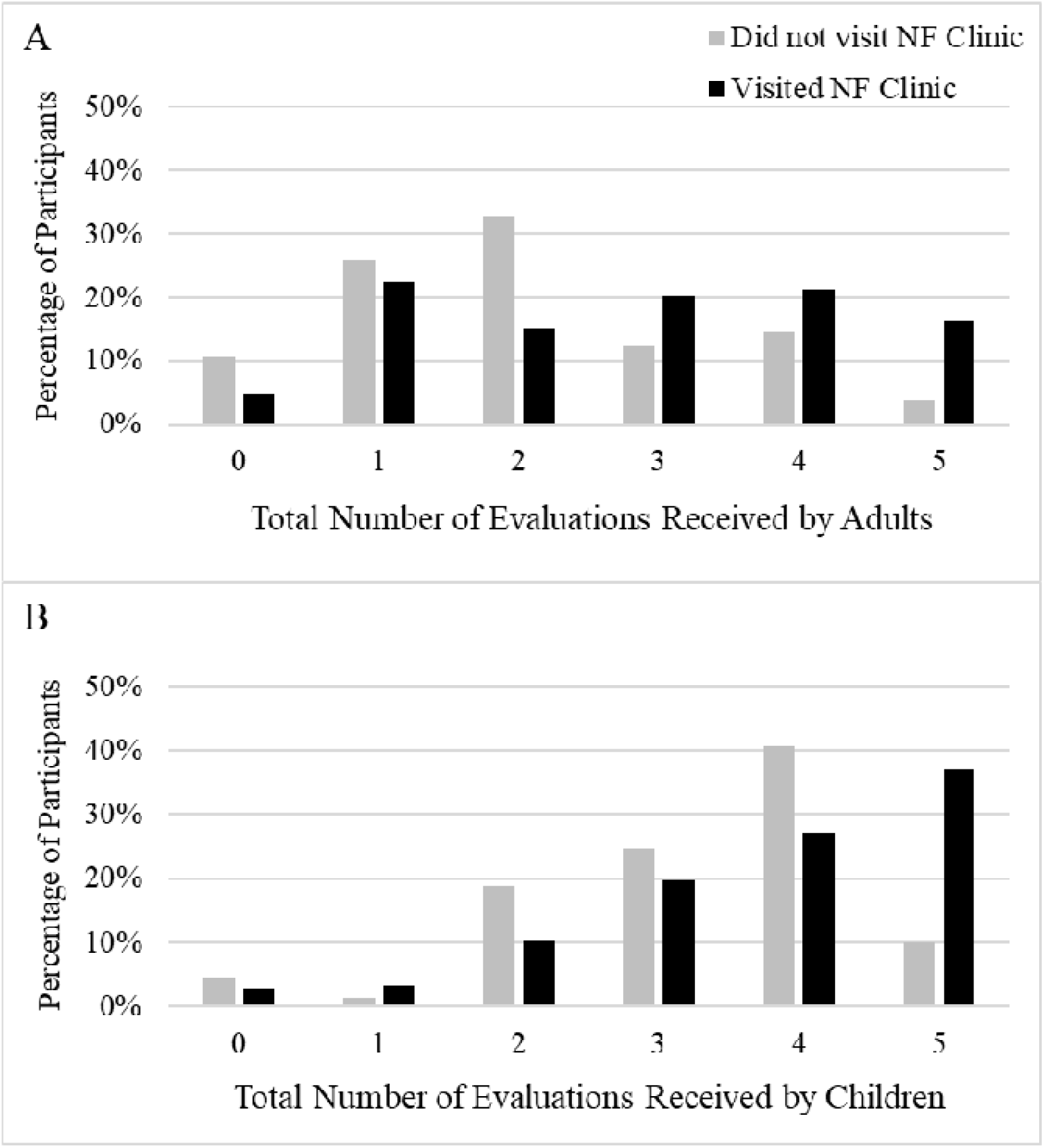
Total Number of Recommended Health Surveillance Evaluations Received by US Adults and Children with NF1 Stratified by NF Clinic Attendance Bars indicate the weighted percentage of NF registry participants estimated to have received the given number of health surveillance evaluations in the prior year. Results are stratified by whether participants had (black) or had not (gray) visited a specialized NF Clinic in the prior 3 years. Panel A: Evaluations assessed for adults included annual blood pressure check, skin exam, scoliosis screening, malignant peripheral nerve sheath tumor education, and family planning education. Panel B: Evaluations assessed for children included annual blood pressure check, skin exam, scoliosis screening, assessment of pubertal development, and tracking of developmental milestones/school progress.

**Table 4.** Factors Affecting Proportion of Health Surveillance Evaluations Received.

| | Children $\beta$ coefficient<br>(p-value) | Adult $\beta$ coefficient<br>(p-value) |
| --- | --- | --- |
| Age (years) | 0.1040 (**p < 0.001**) | -0.016 (**p = 0.03**) |
| Gender |  |  |
| Male | *reference* | *reference* |
| Female | -0.123 (p = 0.55) | 0.096 (p = 0.71) |
| Ancestry |  |  |
| Non-Hispanic White | *reference* | *reference* |
| Black | -0.440 (p = 0.37) | -0.949 (**p = 0.03**) |
| Other<sup>a</sup> | 0.391 (p = 0.11) | -0.700 (**p = 0.04**) |
| Household Income |  |  |
| <\$25,000 | *reference* | *reference* |
| \$25,000-\$129,999 | -0.297 (p = 0.40) | 0.370 (p = 0.25) |
| ≥\$130,000 | -0.283 (p = 0.43) | 0.780 (p = 0.14) |
| Primary Medical Insurance |  |  |
| Medicaid | *reference* | *reference* |
| Medicare | 0.380 (p = 0.54) | 1.346 (**p = 0.008**) |
| Commercial | -0.128 (p = 0.59) | 1.096 (**p = 0.01**) |
| Home Location |  |  |
| Rural | *reference* | *reference* |
| Suburb | -0.176 (p = 0.46) | 0.344 (p = 0.34) |
| Urban | -0.330 (p = 0.32) | 0.135 (p = 0.74) |
| Education |  |  |
| No college degree | N/A | *reference* |
| College degree or higher<sup>b</sup> |  | 0.302 (p = 0.20) |
| NF Specialty Clinic Attendance |  |  |
| No visit within prior 3 years | *reference* | *reference* |
| Visit within prior 3 years | 0.858 (**p < 0.001**) | 0.764 (**p = 0.002**) |
| Last Visit with NF Team |  |  |
| More than one year ago | *reference* | *reference* |
| Within the prior year | 1.616 (**p < 0.001**) | 1.233 (**p < 0.001**) |

Note: Beta ( $\beta$ ) coefficients and p-values were calculated from robust linear regressions adjusted for age. All p-values <0.05 are bolded. <sup>a</sup>Includes Latino/Hispanic, Asian, Native American or Alaskan Native, Native Hawaiian or other Pacific Islander, and Other due to small sample sizes. <sup>b</sup>Includes 2- and 4-year undergraduate degrees and higher.

## Discussion

In this national survey of adults with NF1 and parent/guardians of children with NF1, we explored access, quality, and equity of care for individuals with NF1 across the US, providing important insight into differences between individuals who attend specialized NF clinics and those who do not. The majority of individuals in a large online NF Registry had not been evaluated at a specialty NF clinic in the prior 3 years. Shortly after the publication of NF1 health surveillance guidance by the ACMG and AAP and 14 months into the COVID-19 pandemic, we found variable application of recommended surveillance. Screening and education recommendations were performed in one-third (skin exams for adults) to four-fifths (blood pressure measurement in children) of eligible patients according to patient/parent self-report.

Encouragingly, there were few sociodemographic disparities in care for children with NF1, with access to NF clinics being the primary predictor of receiving recommended health surveillance. However, significant sociodemographic disparities in both access to NF clinics and receipt of recommended NF1 health surveillance were present for adults with NF1. This gap in access to NF clinics likely reflects the paucity of adult NF clinics in the US compared to pediatric NF centers.^5^ As reported in this survey, there are challenges in traveling to, or obtaining insurance coverage for, out-of-state clinics. Limited access to rare disease care, exacerbated by insurance denials for specialist care, is a known problem across many rare diseases,^13,14^ and may be worse for adults due to barriers in transitioning from pediatric to adult centers.^15^ Our findings emphasize the urgent need to develop and evaluate programs and policies that focus on improving care for adults with NF1.

While information on care for US NF1 patients is sparse, and often limited to individuals with specific manifestations of NF1,^16^ research on NF1 care in Australia reinforces and provides additional insight into the limited uptake of recommended health surveillance in our sample. In surveys and interviews with Australian adults with NF1, about half reported not receiving routine health surveillance for NF1, with many individuals unaware of the need for regular monitoring or unsure of where to access care.^17,18^ Qualitative interviews also revealed that some adults saw routine monitoring as unnecessary in the absence of bothersome NF1 symptoms or futile due to the limited treatments for NF1,^18^ echoing similar findings in our survey regarding reasons for not attending NF clinics. Lastly, in qualitative interviews conducted specifically in young adults in Australia, many interviewees had a poor understanding of NF1-associated complications and had not sought care for even severe NF1 symptoms, and this trend was exacerbated by NF1-related cognitive deficits in some individuals.^19^ Overall, this data suggests a need to educate families about NF1 care recommendations, including the benefits of early detection for NF1-associated malignancies as well as education regarding newly available treatment options for non-malignant tumors; education should be delivered in ways that are accessible to individuals with lower educational attainment, lower health literacy, and/or learning disabilities, all of which are common in adults with NF1.^20–22^

Our paper has limitations. While we included responses from a relatively large number of participants with varying engagement in specialty care, our survey had a low response rate. We used inverse propensity scores to attempt to mitigate nonresponse bias in this analysis and are exploring ways to engage a higher proportion of NF registry members in future surveys. Due to difficulties identifying NF1 patients within outpatient claims data^23^ and the inability of medical record reviews to capture care received across health systems, we relied on self-reported data from patients/parents to understand receipt of recommended NF1 care. This data may be subject to inaccurate reporting or recall bias. To limit these biases, medical procedures were described in lay terms in the survey, and our analyses focused on care in the last year. Observed rates of attendance at NF clinics and provision of recommended healthcare may have been temporarily lessened due to the COVID-19 pandemic.^24–26^ To partially address this, we asked individuals whether they had been to an NF clinic in the prior 3 years rather than the prior year, and we plan to track changes in receipt of recommended care through additional surveys over time. Finally, our analysis of factors affecting the receipt of recommended health surveillance used a composite index due to concerns for type 1 statistical error, but the specific barriers inhibiting receipt of each individual recommended assessment may differ. Future research with clinicians from diverse practice settings and specialties must explore the facilitators and barriers to delivering recommended services.

In conclusion, our study demonstrates that a substantial number of NF1 patients in the US do not attend specialized NF clinics and do not receive all NF1-related health surveillance recommendations. Promoting patient awareness of specialized clinics, utilizing telemedicine and hub-and-spoke models of care, and insurance reform are strategies that may improve access to NF clinics. However, given currently limited access to specialty NF clinics, efforts to develop and implement NF care guidelines should also prioritize broad dissemination to non-disease specialists. To do this successfully, increased stakeholder engagement and a focus on the ease of implementation of guidelines in everyday clinical practice is essential.^27^ Furthermore, disseminating lay language explanations of NF1 care recommendations directly to people with NF1 and their family members could empower them to seek indicated medical services, as has been demonstrated in other genetic disorders such as trisomy 21.^28^ Together, these initiatives may reduce disparities in NF1 care in the US, ensuring all NF1 patients have access to high-quality care for their condition.

## Data Availability

All data may be requested from the NF Registry by emailing. All requests are reviewed by the NF Registry Data Access Committee to ensure appropriate scientific and ethical use of the data.

## Acknowledgments

The authors would like to thank Pamela Knight, MS, for assistance with data collection through the NF Registry and Liesel Von Imhof, BA, for assistance with cleaning the data for analysis.

## Author Contributions

Conceptualization: V.M., H.R., K.Y., N.U., S.P., J.J.; Data curation: K.K.; Formal analysis: V.M., YM., L.C.; Methodology: V.M., Y.M., L.C., S.P., J.J.; Resources: H.R., K.K.; Software: Y.M., L.C.; Supervision: S.P., J.J.; Visualization: V.M., Y.M..: Writing-original draft: V.M.; Writing-review & editing: Y.M., L.C., H.R., K.K., K.Y., N.U., S.P., J.J.

## Appendix. Online-Only Supplemental Tables

**Supplemental Table 1.**
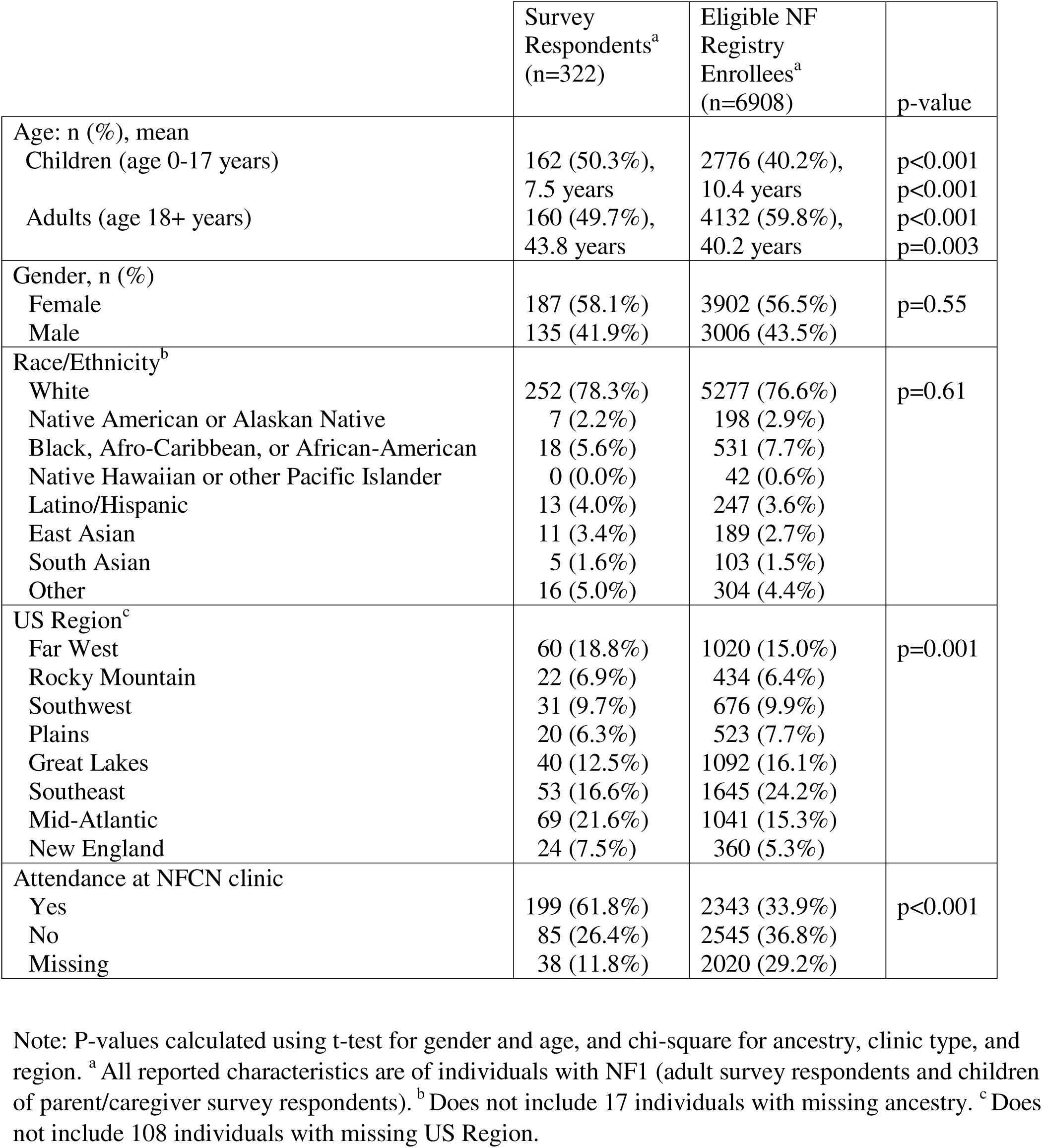
Characteristics of Survey Respondents and Eligible NF Registry Enrollees.

|  | Survey Respondents<sup>a</sup><br>(n=322) | Eligible NF Registry Enrollees<sup>a</sup><br>(n=6908) | p-value |
| --- | --- | --- | --- |
| Age: n (%), mean |  |  |  |
| Children (age 0-17 years) | 162 (50.3%),<br>7.5 years | 2776 (40.2%),<br>10.4 years | p<0.001<br>p<0.001 |
| Adults (age 18+ years) | 160 (49.7%),<br>43.8 years | 4132 (59.8%),<br>40.2 years | p<0.001<br>p=0.003 |
| Gender, n (%) |  |  |  |
| Female | 187 (58.1%) | 3902 (56.5%) | p=0.55 |
| Male | 135 (41.9%) | 3006 (43.5%) |  |
| Race/Ethnicity<sup>b</sup> |  |  |  |
| White | 252 (78.3%) | 5277 (76.6%) | p=0.61 |
| Native American or Alaskan Native | 7 (2.2%) | 198 (2.9%) |  |
| Black, Afro-Caribbean, or African-American | 18 (5.6%) | 531 (7.7%) |  |
| Native Hawaiian or other Pacific Islander | 0 (0.0%) | 42 (0.6%) |  |
| Latino/Hispanic | 13 (4.0%) | 247 (3.6%) |  |
| East Asian | 11 (3.4%) | 189 (2.7%) |  |
| South Asian | 5 (1.6%) | 103 (1.5%) |  |
| Other | 16 (5.0%) | 304 (4.4%) |  |
| US Region<sup>c</sup> |  |  |  |
| Far West | 60 (18.8%) | 1020 (15.0%) | p=0.001 |
| Rocky Mountain | 22 (6.9%) | 434 (6.4%) |  |
| Southwest | 31 (9.7%) | 676 (9.9%) |  |
| Plains | 20 (6.3%) | 523 (7.7%) |  |
| Great Lakes | 40 (12.5%) | 1092 (16.1%) |  |
| Southeast | 53 (16.6%) | 1645 (24.2%) |  |
| Mid-Atlantic | 69 (21.6%) | 1041 (15.3%) |  |
| New England | 24 (7.5%) | 360 (5.3%) |  |
| Attendance at NFCN clinic |  |  |  |
| Yes | 199 (61.8%) | 2343 (33.9%) | p<0.001 |
| No | 85 (26.4%) | 2545 (36.8%) |  |
| Missing | 38 (11.8%) | 2020 (29.2%) |  |

Note: P-values calculated using t-test for gender and age, and chi-square for ancestry, clinic type, and region. <sup>a</sup> All reported characteristics are of individuals with NF1 (adult survey respondents and children of parent/caregiver survey respondents). <sup>b</sup> Does not include 17 individuals with missing ancestry. <sup>c</sup> Does not include 108 individuals with missing US Region.

**Supplemental Table 2.** Self-reported Reasons for Not Seeking Care at a Specialized NF Clinic.

|  | Adults with NF1 |  |  |  | Parents of Children with NF1 |  |  |  |
| --- | --- | --- | --- | --- | --- | --- | --- | --- |
|  | Major Problem | Minor Problem | Not a Problem | Missing | Major Problem | Minor Problem | Not A Problem | Missing |
| Insurance doesn't cover specialized clinics or they are out of network | 19% | 21% | 45% | 15% | 7% | 27% | 57% | 9% |
| Didn't know there were specialized NF clinics | 18% | 22% | 45% | 15% | 17% | 17% | 58% | 9% |
| Don't want to travel to specialized NF Clinic/ too far away | 15% | 24% | 46% | 15% | 8% | 56% | 28% | 7% |
| Can't travel to specialized NF clinic (due to my health, cost of travel, etc) | 15% | 14% | 56% | 15% | 11% | 15% | 66% | 9% |
| Don't see the value in attending a specialized NF clinic at this time | 6% | 17% | 62% | 15% | 4% | 35% | 52% | 9% |

Note: All percentages are weighted using inverse propensity scores based on age, region, and NFCN clinic attendance within responses from 88 adults and 45 parents of children with NF1 who did not attend or were unsure whether they attended a specialized NF clinic in the prior 3 years.

